# TEG Max Clot Strength is Consistently Elevated and May Be Predictive of COVID-19 Status at the Time of ICU Admission

**DOI:** 10.1101/2020.04.30.20076703

**Authors:** Shaun Lawicki, Katherine Wang, Bing Han, Gordon Love

## Abstract

**Background:** Hypercoagulability is becoming widely recognized as a major complication of COVID-19 infection as evidenced by high levels of fibrinogen degradation products and microthrombi identified within the lungs and kidneys of autopsy specimens from these patients. We report thromboelastography (TEG) testing on a cohort of patients with suspected COVID-19 infection at the time of admission to the intensive care unit.

**Methods:** TEG testing was performed using the TEG 6s analyzer near or at the time of ICU admission. We also report the results of other coagulation or inflammatory related indices such as platelet count, prothrombin time, fibrinogen, D-dimer, C-reactive protein, ferritin, and procalcitonin. All laboratory testing was performed at the discretion of the attending physician in the course of normal patient care and retrospectively reviewed.

**Results:** We found that maximum clot strength was consistently elevated in COVID-19 patients while normal in all patients found to be negative. We did not encounter significant prolongations of coagulation assays outside of those expectedly prolonged by heparin therapy nor was meeting the criteria for disseminated intravascular coagulation encountered.

**Conclusions:** We postulate that elevated maximum clot strength by TEG testing is predictive of COVID-19 status as within our cohort this perfectly predicted patients’ COVID-19 status despite a high level of suspicion in negative patients with normal TEG results. While these results require a larger cohort for confirmation, we feel that TEG testing could improve confidence in COVID-19 testing results in suspected patients possibly allowing for earlier de-escalation of infectious precautions and personal protective equipment utilization.

## Introduction

A large part of the dialogue on COVID-19 is focused on an association between COVID-19 and hypercoagulability. It has been reported that many severe COVID-19 patients meet criteria for disseminated intravascular coagulation (DIC).^1^ A summary of Tang et al.’s report showed that 71.4% of patients who did not survive the infection were reported to have developed DIC by day 4, as compared to just one patient (0.6%) who developed DIC in the group that survived.^2^ That being said, it should be kept in mind that the International Society for Thrombosis and Hemostasis (ISTH) criteria for DIC is validated for use in conditions known to be associated with DIC which has not yet been proven in COVID-19. It is also worth noting that these criteria are largely met in this study based on the markedly elevated D-dimer levels found in COVID-19 patients who had worse outcomes.^1^ However, D-dimer may be a product of local thrombin formation as well as through a disseminated process. A series of COVID-19 autopsy findings from our institution would support this theory. In our autopsy cases, fibrin thrombi were found to be present within the capillaries and small vessels of the lung. Moreover, CD61+ megakaryocytes were observed in association with heightened platelet generation and subsequent aggregation into platelet-rich thrombi. Of additional note, these megakaryocytes displayed significant nuclear hyperchromasia and atypia, raising the question of direct viral infection of these cells, which was seen in severe SARS.^3^

Undoubtedly, COVID-19 is associated with a pro-thrombotic state. It has been advocated that multiple parameters, including D-dimer, platelet count, PT, and fibrinogen values be followed to monitor for coagulopathy.^2^ However, currently published literature has not looked at the utility of thromboelastography (TEG) testing in COVID-19 patients. Following discussion of the common findings in our local institution’s autopsy series, we decided to review coagulation and inflammatory testing performed in our COVID-19 patient cohort. We report the first series of TEG findings in suspected COVID-19 patients at the time of ICU admission.

## Methods

All patients reviewed were admitted to one of the intensive care units at University Medical Center New Orleans (UMCNO) with suspicion for COVID-19 infection. All laboratory testing was performed at the discretion of the attending physician as part of normal patient care. TEG testing was performed by the UMCNO blood bank using TEG 6s analyzers with either citrated: Kaolin, Rapid TEG, Functional Fibrinogen (which include the LY30 parameter) or citrated: Kaolin, Kaolin Heparinase, Rapid TEG, Functional Fibrinogen (which do not include LY30) cartridges (Haemonetics, Braintree, MA). All other laboratory testing was performed in the UMCNO core laboratory per the manufacturers’ instructions for each assay. Coagulation testing was performed on ACL Top 700 analyzers (Instrumentation Laboratory, Bedford, MA). CBC indices, C-reactive protein (CRP), and ferritin were performed on DxH 900, AU5800, and Unicel Dxl 600 analyzers respectively (Beckman Coulter, Brea, CA). Procalcitonin was performed on a VIDAS 3 analyzer (bioMérieux, Marcy-I’Etoile, France).

TEG testing was generally performed at the time of ICU admission often before the patient’s COVID-19 status was known. We report the results of all other testing which was most closely associated chronologically with the time of TEG testing. This study received an institutional review board (IRB) exemption from the Louisiana State University Health Science Center New Orleans IRB and approval from the Research Review Committee of UMCNO.

## Results

All patients were adults with an average age of 61.9 years, 46% were male, and 76.9% were African American. Co-morbidities were common with 30.8% having cardiac disease, 46.2% with diabetes, 15.4% with pulmonary disease, 26.9% with chronic kidney disease, 30.8% with hyperlipidemia, and 76.9% with hypertension (Table 1). There is 38.5% mortality in our cohort with 30.8% discharged and 30.8% still admitted. The range of follow-up from ICU admission is 2–28 days. 42.6% of patients were anticoagulated with continuous IV heparin at the time of TEG testing due to evidence of acute kidney injury. All other patients received anticoagulation with prophylactic dose Lovenox. 80.8% of our patients were found to be infected with COVID-1 9.

**Table 1.**
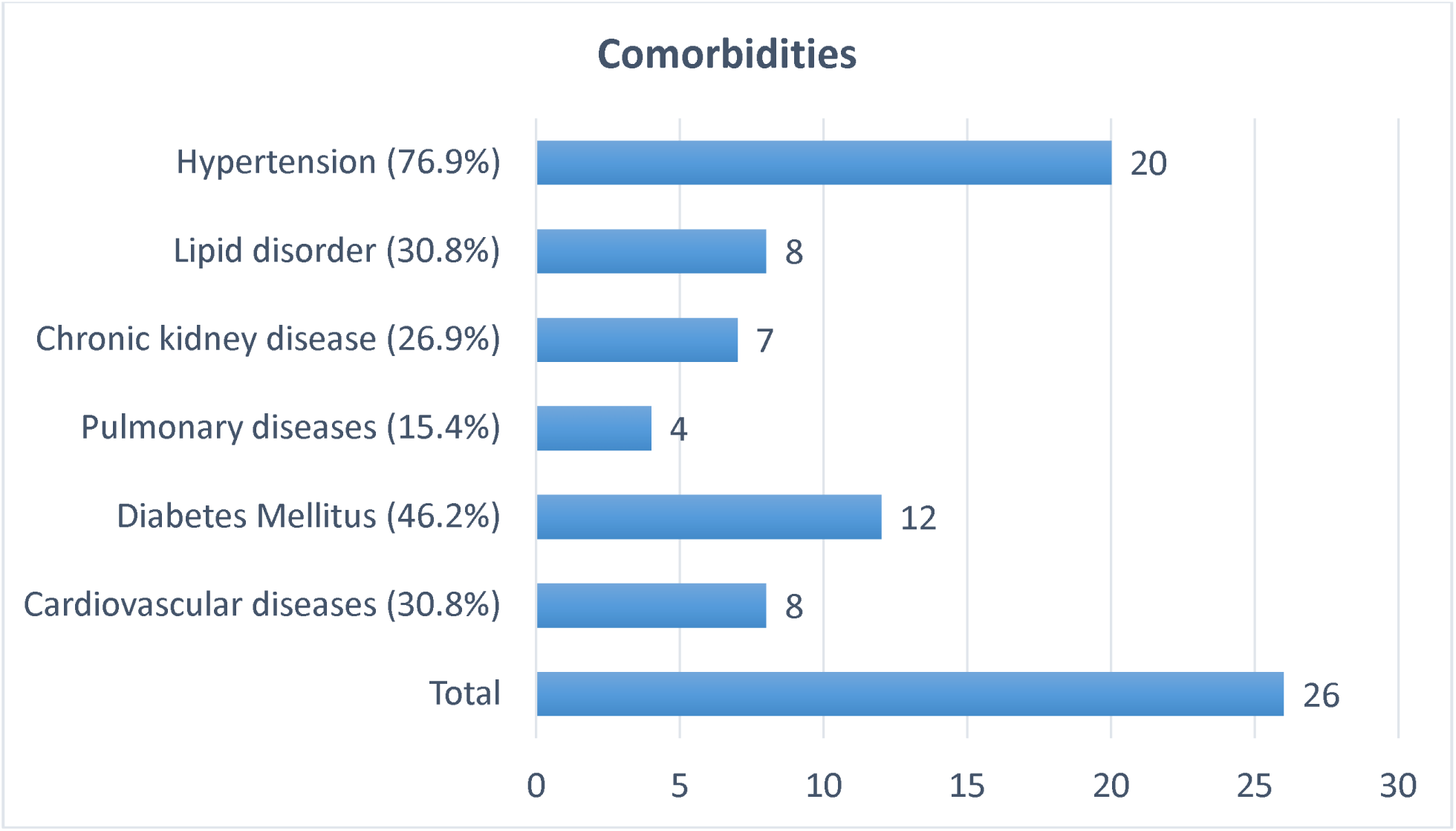
Comorbidities present in our cohort of patients.

We found that TEG testing revealed a consistently elevated functional fibrinogen maximum clot strength in all COVID-19 positive patients and an elevated rapid TEG maximum clot strength in all but two COVID-19 positive patients. These two patients still showed high normal rapid TEG maximum clot strength despite moderate thrombocytopenia in one patient and daily aspirin therapy in the other. Clot times were prolonged in all patients receiving IV heparin at the time of testing as expected but were normal in non-heparinized patients or following heparin neutralization with heparinase. All patients who were found to be COVID-19 negative had normal TEG results (Table 2).

**Table 2.**
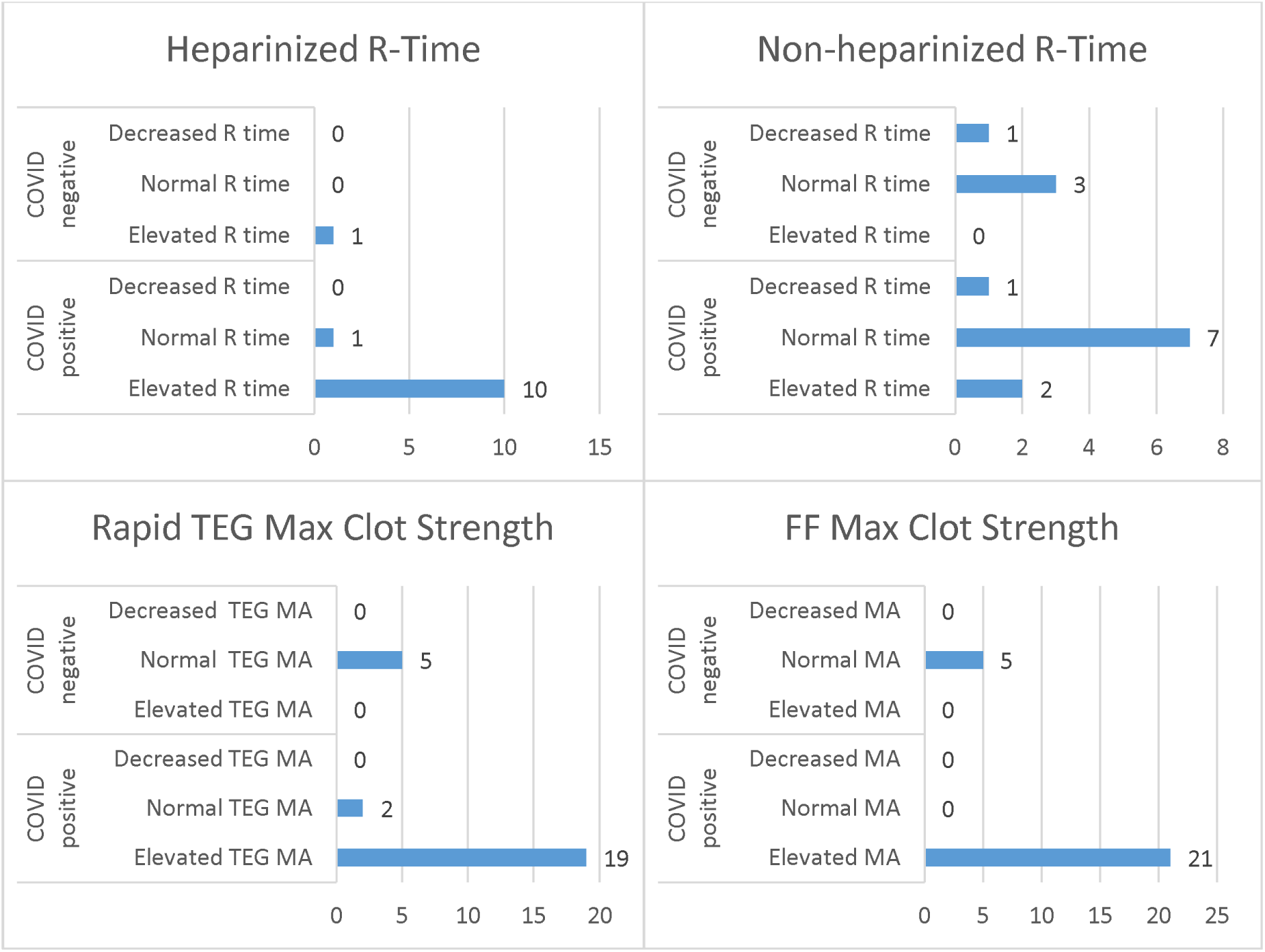
TEG results divided into COVID-19 positive and negative patient groups.

Only one of our patients (3.8%) was thrombocytopenic at the time of testing and none had significant abnormalities of PT/INR. Fibrinogen testing was consistently elevated in COVID-19 positive patients when performed. D-dimer was commonly but not consistently elevated (22/24 cases, 91.7%). Other markers of acute inflammation such as CRP (22/24 cases, 91.7%), ferritin (19/25 cases, 76%), and procalcitonin (16/20 cases, 80%) were also commonly but not consistently elevated (Table 3). It is also noteworthy that elevations of these markers were also common in our five COVID-19 negative patients with 80% having elevated D-dimer, 60% having elevated procalcitonin and CRP, and 40% having elevated ferritin.

**Table 3.**
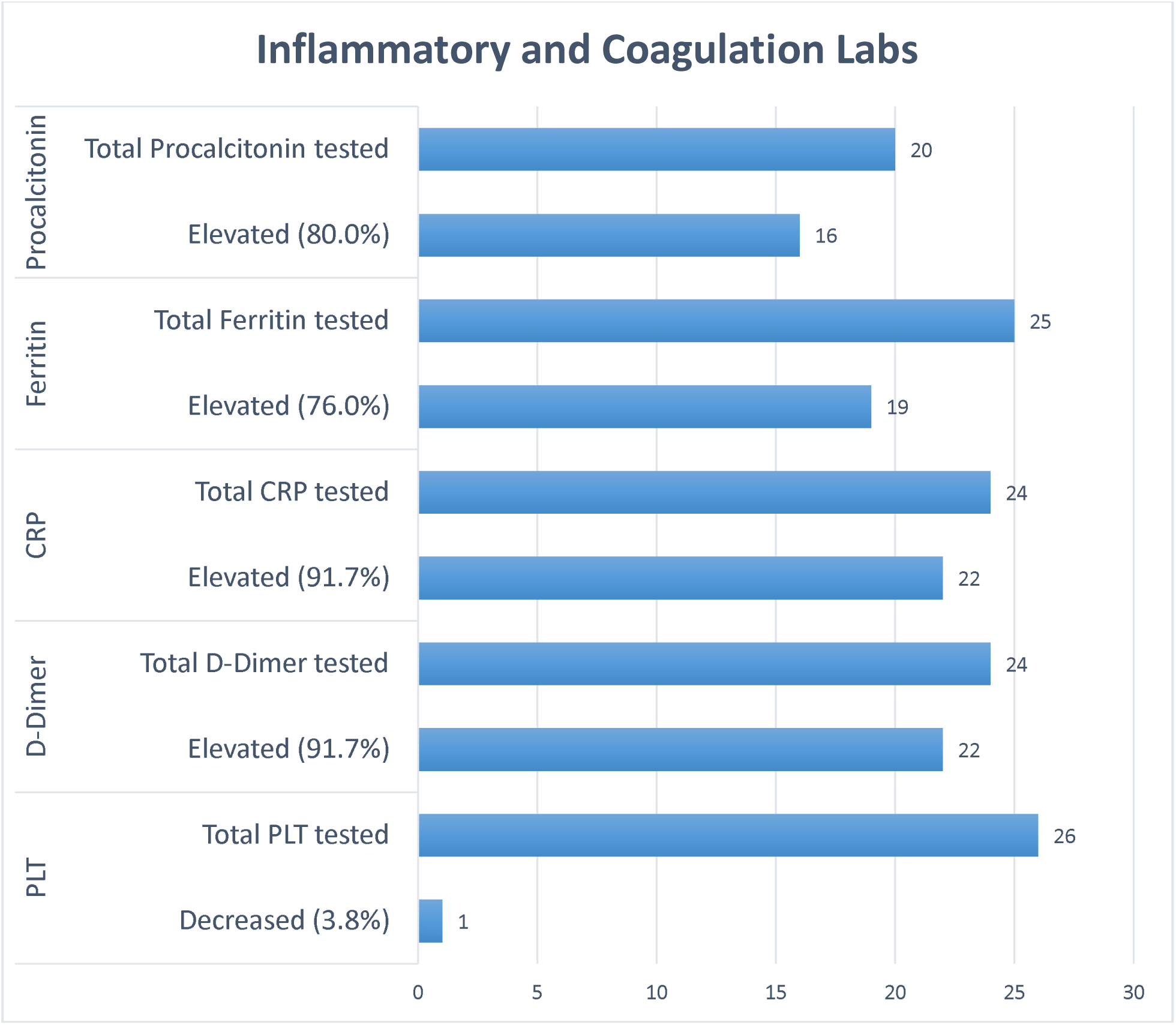
Results of selected laboratory tests in our patient cohort.

None of our cohort met the ISTH criteria for DIC over his or her hospital course although only a minority of patients (19.2%) in our cohort had all laboratory testing evaluated by the ISTH criteria as many patients did not have fibrinogen results available. However, we note that even if fibrinogen was low in these patients, they still would not have met criteria for DIC, and this would appear to be an unexpected result as fibrinogen was consistently elevated in our cohort when performed.

## Discussion

Hypercoagulability is quickly being recognized as a potentially serious complication of COVID-19 infection as evidenced by the common finding of platelet rich microthrombi identified in our institution’s autopsy series.^3^ A high rate of venous thromboembolism has also been reported.^4^ The likely contribution of this hypercoagulability to illness severity and mortality is also highlighted by the report of a potential survival benefit in COVID-19 patients who receive anticoagulation with heparin.^5^ While TEG testing is not commonly performed in ICU patients and may not be widely available, we report the first series of results from COVID-19 patients utilizing this assay given its increased clinical utility over standard coagulation assays such as PT/INR and PTT which are known to not be particularly predictive of a pro-thrombotic state.

While it has been reported that meeting the ISTH criteria for DIC is common in non-survivors of COVID-19, we did not find that this was common in our patient cohort. It is our opinion that DIC may be a misnomer for the abnormal coagulation present in these COVID-19 patients except possibly as an end-stage event near the end of life in the most severely afflicted although even non-survivors in our cohort never met the ISTH criteria. Since evidence of unchecked thrombin generation has not yet been shown in COVID-19 patients and subsequent coagulation factor and platelet consumption is not a prominent feature as in most other cases of DIC, we feel these patients are more consistent with a hypercoagulability likely secondary to activation from crosstalk between the inflammatory and coagulation systems as well as local activation in the lung due to or in concert with the acute lung injury present with this disease. However, we recognize this opinion remains conjecture until more robust studies of the underlying mechanisms are conducted.

Our retrospective study found that COVID-19 positive patients have consistently elevated functional fibrinogen and rapid TEG maximum clot strengths while COVID-19 negative patients had normal TEG results. Therefore, we propose that elevated maximum clot strengths by TEG may be predictive of COVID-19 status in patients at the time of ICU admission as we found in our cohort of patients. Of note, this held true even in two patients where COVID-19 suspicion was high enough to suspect a false negative initial test and upon retesting were still found to be negative. This potentially predictive nature was not present for other markers of inflammation or coagulation as there were also commonly elevated values in COVID-19 negative patients and occasional normal results in the COVID-19 positive group. Although our results could prove useful for limiting unnecessary retesting and possibly earlier de-escalation of infectious precautions with subsequent saving on precious personal protective equipment, we recognize that our findings have several limitations. First, our sample size is small especially our group of COVID-19 negative patients making it difficult to predict how consistent elevated maximum clot strength may be predictive in a larger cohort or at a different time in patients’ hospital courses since all our patients were admitted to the ICU at the time of TEG testing. It is noteworthy that one of our COVID-19 positive patients did receive TEG testing at the time of original presentation over a week before ICU admission when symptoms were still mild-moderate, and this was normal. Second, we have not been able to perform more esoteric coagulation studies in these patients, so we are unable to do anything other than speculate on the underlying mechanism of these elevated maximum clot strengths. Since both functional fibrinogen and rapid TEG maximum clot strengths are elevated it is unknown if these abnormalities are due to increased platelet function/activation, elevated fibrinogen production, increased thrombin generation (locally in the lung or systemically), or a combination of all three. It has also been reported that the TEG functional fibrinogen assay does not eliminate the platelet contribution to maximum clot strength completely.^6^ Finally, TEG results where available are generally performed expediently with results available within 1–2 hours which at the time of this study was significantly faster than COVID-19 testing results, however we recognize that COVID-19 testing capabilities are rapidly expanding and may now be available within a similar time frame at some institutions. We suggest that performing these tests in parallel could still improve confidence in the COVID-19 result and potentially lower repeat testing.

## Conclusion

We report the first series of TEG results in suspected COVID-19 patients and its potential to be predictive of COVID-19 status at the time of ICU admission. A more complete understanding of the mechanisms of the apparent hypercoagulability present in severe COVID-19 infection remains to be elucidated. We feel that additional studies into platelet function/activation, thrombin generation, and natural anticoagulant factors in COVID-19 patients are needed along with TEG testing in COVID-19 patients at different time points over the disease course which may also reveal a prognostic role for TEG.

## Data Availability

No additional or supplemental datasets are available in association with this manuscript

## Acknowledgements

We would like to acknowledge our clinical colleagues for the initial consideration for performing TEG testing, and putting themselves at risk caring for our COVID-19 patients. We also appreciate the ongoing dedication of our blood bank and laboratory staff in this difficult time.

## Author Contribution

SL, KW, BH contributed to the data collection and review as well as the manuscript writing and editing. GL provided general oversight and manuscript review.

